# Dose-response modelling of endemic coronavirus and SARS-CoV-2: human challenge trials reveal the individual variation in susceptibility

**DOI:** 10.1101/2022.04.07.22273549

**Authors:** Fuminari Miura, Don Klinkenberg, Jacco Wallinga

**Author notes:** **Corresponding author** F. Miura.

## Abstract

We propose a mathematical framework to analyze and interpret the outcomes of human challenge trials. We present plausible infection risks with HCoV-229E and SARS-CoV-2 over a wide range of infectious dose, and suggest ways to improve the design of future trials and to translate its outcomes to the general population.

**One sentence summary:** We rephrase dose-response models in terms of heterogeneity in susceptibility in order to present the possible range of infection risks for endemic coronaviruses and SARS-CoV-2

## Background

Quantifying the infectivity of pathogens is a crucial step towards the understanding of infection risks. In human challenge trials the infection risk is observed as the proportion of exposed participants that become infected. Dose-response models describe how this proportion infected changes with an increase in the infectious dose used to expose the participant [1,2]. Such dose-response models can be used to improve trial designs [3], to describe infectivity and immunogenicity in human hosts [4], and to simulate the infection risks via various transmission routes [5].

Dose-response models can account for variation in host susceptibility, and most often such variation has been modelled by a beta distribution [6,7]. However, when a proportion of individuals is completely immune, the variation is better captured by other distributions (e.g., [8]).

Here, we start by reformulating dose-response models with a flexible description of the variation in host susceptibility that allows for an intuitive biological interpretation. We show how variation in susceptibility determines the dose-response relationship for the endemic human coronavirus HCoV-229E and we compute the plausible range of SARS-CoV-2 dose-response curves based on available outcomes of a challenge study.

Our approach suggests how the design of human challenge trials can be improved to better capture the variation in susceptibility, and suggests how to translate the outcomes of human challenge studies into infection risks for the general population.

## Methods

### Human challenge studies

We conducted a literature search to collect available data from human challenge studies with endemic coronaviruses and SARS-CoV-2. The collected data consists of 5 studies with endemic coronaviruses HCoV 229E and one study with SARS-CoV-2. In all cases the study population consisted of healthy adult volunteers, and the participants were intranasally inoculated with certain doses in each trial. The challenge studies reported the challenge dose, the number of challenged individuals, and the number of infected individuals as summarized in **Supplementary Material, Table S1** and **Table S2**.

### Dose-response models to analyze human challenge studies

The reported infectious doses *d* are expectations of a Poisson distribution of the actual infectious dose, with mean *d*. If each host is equally susceptible, the probability of infection *P*(*d*) given a challenge dose *d* is *P*(*d*) = 1 − *exp*(−*d*). This assumes that each infectious particle can independently establish an infection [2,9].

We capture the variation in susceptibility among study participants by assigning each a level of susceptibility *s*, according to a distribution *f*(*s*) with mean 1. A participant with a level of susceptibility *s* has *s* times higher probability of infection compared to an average individual, per infectious particle. The probability of infection upon challenge with a dose *d* is 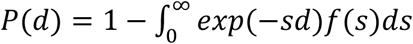. By fitting this model for the probability of infection to the observed proportion of infections in human challenge studies we can infer the shape of the distribution *f*(*s*) using the method of maximum likelihood, see **Supplementary Material** for details. We use four different models for the distribution: a Dirac delta distribution to reflect a situation where all individuals have the same level of susceptibility; a gamma distribution to reflect a situation where the level of susceptibility varies continuously; a bimodal distribution with one fraction of the population almost immune, and the remaining fraction with a single level of susceptibility; and a bimodal distribution with one fraction of the population almost immune, and the remaining fraction with a gamma distribution for the level of susceptibility to vary continuously. Detailed model descriptions and estimated parameters are provided in **Table S3**.

## Results

### Susceptibility distributions determine the shape of dose-response curves

We described the proportion of infections among individuals exposed to different doses of the endemic coronavirus HCoV-229E by fitting dose-response models. Since the collected trial data include participants who might have been exposed to viruses, we included bimodal distributions of susceptibility.

The results reveal a strong statistical support for a distribution reflecting a situation where a fraction of the population is almost immune whereas the remaining fraction of the population has a single level of susceptibility. There is no statistical support for a homogeneous level of susceptibility or continuous variation in susceptibility for all individuals (**Table S4**).

### Plausible SARS-CoV-2 dose-response curves

In the available human challenge study with SARS-CoV-2 all participants were healthy young adults with no evidence of previous SARS-CoV-2 infection or vaccination, and they were all exposed to the same single dose [10]. Here we show how the variation in susceptibility would affect the infection risk at different doses. Since participants had not had any prior exposure to this virus, we assumed a continuous variation in susceptibility only. We fitted the model with several gamma distributions for the level of susceptibility *s* to the observed SARS-CoV-2 challenge data, where we increased the coefficient of variation (*CV*) over orders of magnitude from small (10^−6^) to large (10^2^). The corresponding curves reveal that the infection risk increases more gradually with increasing CV in susceptibility (**Figure 1A**).

**Figure 1.**
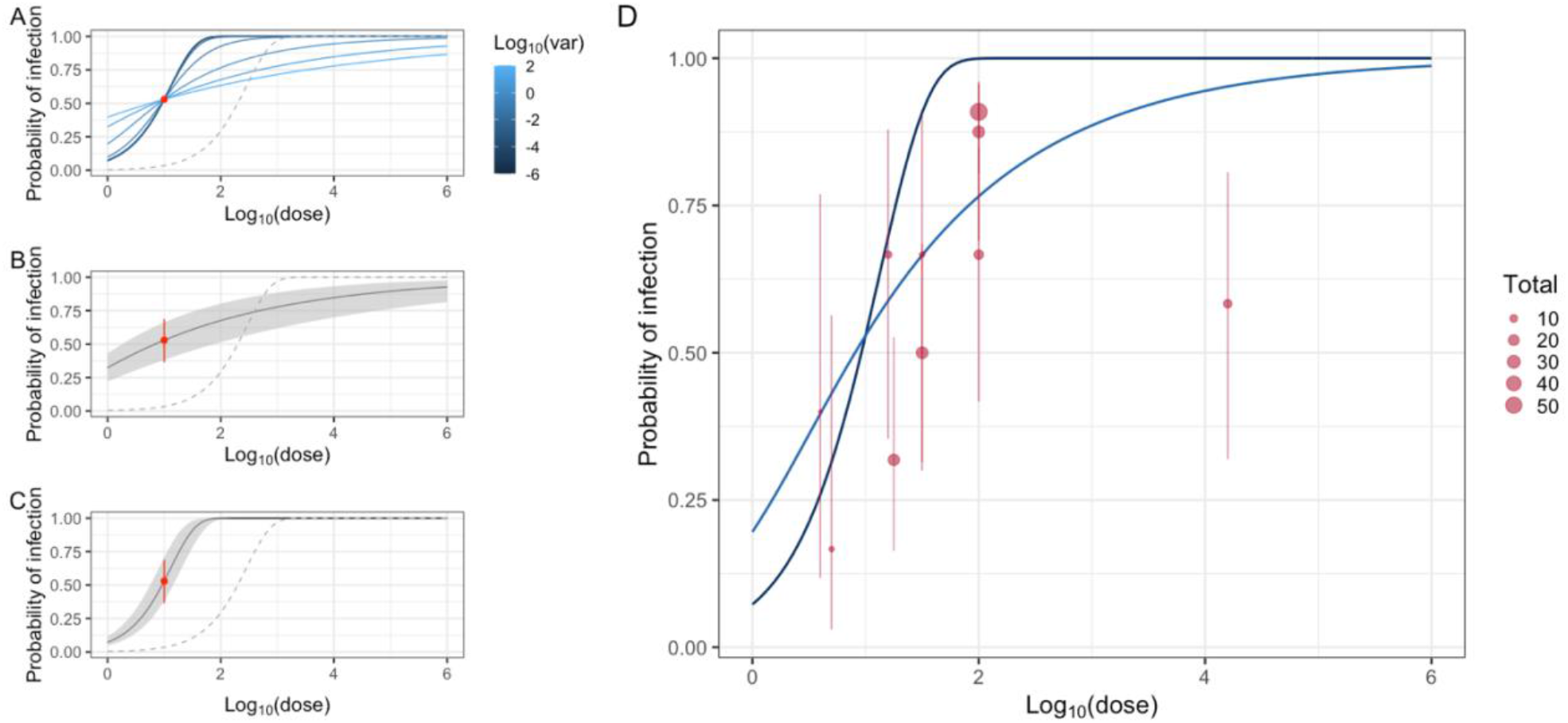
Dose-response curves of SARS-CoV-2 (panel **A, B**, and **C**) and comparison with an estimated dose-response curve of endemic coronaviruses (panel **D**) based on observed human challenge data. In panel **A**-**C**, light red dots stand for observations from human challenge data of SARS-CoV-2 (**Table S2**), and whiskers show Jeffrey’s binomial confidence intervals (95%). Panel **A** illustrates that the steepness of fitted curves (blue lines) decreases over the increase in the coefficient of variation (CV) in susceptibility from 10^−6^ to 10^2^. Panel **B** and **C** show curves fitted to 1000 bootstrapped samples from observed data where the CV is set as 0 and 1 respectively. These values are selected to consider two scenarios where the susceptibility level is completely homogeneous and where the level is comparable to that of endemic coronavirus infection. Dotted grey lines show reference dose-response curves of SARS-CoV-1 animal model [11]. In panel **D**, plots and whiskers are observed challenge data of endemic coronaviruses (**Table S1**), and dark and light blue lines indicate fitted SARS-CoV-2 models where the CV is fixed as 0 and 1 respectively.

We accounted for the statistical uncertainty in the fraction of the participants that were infected by taking 1000 bootstrap samples and fitting dose-response curves to each bootstrapped dataset, reflecting a situation where the susceptibility level is completely homogeneous (*CV* = 0), and where the level is similar to that of the endemic coronavirus infection (*CV* = 1)(**Figure 1B** and **1C**).

We compared the bootstrapped SARS-CoV-2 dose-response curves with the dose-response curve from a SARS-CoV-1 mouse model obtained by Watanabe et al. [11], which has been widely used in risk assessments of SARS-CoV-2 (dotted lines in **Figure 1**). This reveals that using the current reference model based on mouse data with SARS-CoV-1 could lead to serious underestimation of the infection risk for SARS-CoV-2, irrespective of the shape of the distribution of susceptibility level. We also compared the SARS-CoV-2 dose-response curves with the observed outcomes from the challenge studies with endemic coronavirus (**Figure 1D**). This suggests that the estimated range of infection risks of endemic coronavirus is consistent with the observed infection risk in the SARS-CoV-2 trial [12].

## Discussion

In this study, we revealed the plausible range of infection risk over multiple orders of magnitude of the infectious dose for the endemic coronavirus HCoV 229E and SARS-CoV-2, based on human challenge trials. We presented how these dose-response relationships are shaped by the underlying distribution of susceptibility to infection.

The range of SARS-CoV-2 dose-response curves arises from the unknown distribution of background susceptibility in the population and the statistical uncertainty due to the limited number of participants that have participated in the human challenge study. Our results caution against assuming equal susceptibility in the population in risk assessments [5,11], as this assumption results in a lower bound for infection risks at lower doses.

Our results provide implications for further research. We address three of them here.

Firstly, our approach suggests a possible improvement in the design of human challenge trials. Conventional trials tend to use a single dose such as the median human infectious dose (HID_50_) [13]. Using two (or more) challenge doses would be highly informative for extrapolating the findings over a wider range of doses. If multiple challenge doses would not be feasible, we advise to consider using a different challenge dose than the doses used in previous studies. The outcomes of the different studies can be combined in a meta-analysis that takes advantage of those different doses to infer how infection risk changes with dose. This would elucidate the unknown variation in level of susceptibility among individuals.

Secondly, the dose-response models proposed here, as many other dose-response models, have underlying assumptions that the infectious particles are homogeneously mixed in the inoculates and act independently in causing an infection [2,9]. These assumptions suffice for describing the outcome of human challenge studies, even though it might not hold, for example, when virus particles aggregate. The dose-response model can be extended to allow for variation in the per-particle probability, using methods explored previously [2,14], which would allow for a built-in check of violating this assumption.

Thirdly, our approach offers guidance for the translation of the observed risk of infection in human challenge studies to the general population. Currently this translation is difficult as the study population of challenge studies, due to safety and ethical reasons, consists of healthy adult volunteers [15]. Such a study population is not representative of a general population which includes children and elderly. Besides, the general population now includes persons who have been exposed to SARS-CoV-2. The strict selection of healthy adult study participants with no evidence of previous exposure inevitably reduces the variation in susceptibility relative to the general population. The proposed dose-response models with flexible distribution of susceptibility allows for exploring the impact of an expected increase in variability of the level of susceptibility in the general population and a flattening of the dose-response curve.

In conclusion, our study reveals plausible dose-response relationships for SARS-CoV-2, based on information from human challenge trials, that are consistent with dose-response curves obtained for human endemic coronaviruses. Human challenge trials would be more informative if they use different doses. When translating the observed infection risks in the specific study population to the general population, the expected higher variability of susceptibility in the general population should be taken into account.

## Data Availability

All codes and analyzed data are available online at the GitHub link (https://github.com/fmiura/CoronaDR_2022).

## Funding

FM acknowledge funding support from JSPS KAKENHI (Grant number 20J00793). This project has received funding from the European Union’s Horizon 2020 research and innovation programme - project EpiPose (JW and DK, Grant agreement number 101003688). This work reflects only the authors’ view. The European Commission is not responsible for any use that may be made of the information it contains.

## Competing interests

The authors declare no competing interests.

## Data and materials availability

all codes and analyzed data are available at the author’s GitHub link (https://github.com/fmiura/CoronaDR_2022).

## Supplementary Materials for

### Materials and Methods

#### Human challenge data

The challenge data of endemic coronaviruses and SARS-CoV-2 were collected from published articles. We conducted literature search using PubMed and Google Scholar, and 13 human challenge studies were found in total. For further analysis, 7 studies [1–7] were excluded because the information on inoculated doses was unavailable. Thus, 5 studies of endemic coronaviruses [8–12] and 1 study of SARS-CoV-2 [13] were used in the dose-response analysis. The data consisted of the number of exposed doses, total participants, and infected individuals in each trial. The summary of analyzed data with details (i.e., inoculation methods and references) is shown in **Table S1**.

To synthesize the obtained data, we set two assumptions. First, the infection status was comparable across those studies. Several studies defined the infection status by antibody level, while others defined it by the presence of viruses. Second, there was negligible effect of aggregation of viruses. Since the detailed information of inoculated samples was not available, the unit of dose is defined as the reported unit (i.e., TCID_50_, Median tissue culture infectious dose). If there is data that quantify the level of aggregation, further extension of the dose-response analysis is also possible (see [14,15]).

#### Rephrased dose-response model

Here we denote the probability of infection in controlled infection experiments as *P*_*inf*_(*d*), a function of dose *d*. In a host, it is reasonable to assume that all the particles are independently infectious and effective to establish an infection (i.e., single-hit theory [15,16]). The simplest dose-response relationship is formulated by incorporating Poisson uncertainty in a microbial inoculum:

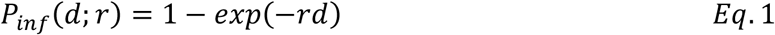

where *r* is the probability of establishing infection by a single-hit.

While previous studies formulated the variation in *r* (often with a beta distribution [15,17–19]), here we focus on the variation in a host. Suppose that the susceptibility to infection among individuals differs and is distributed as *f*(*s*) with a level of susceptibility *s*. The interpretation of variable *s* is that an individual with the level of susceptibility *s* = *s*′ has *s*′ times higher probability of infection compared to an individual with *s* = 1. By expanding *Eq*.1 and integrating the variation in susceptibility, the marginal probability of infection is written as

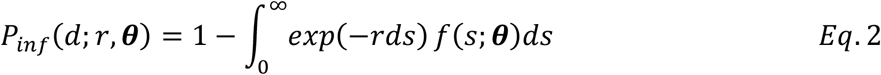

where ***θ*** is a parameter vector of *f*(*s*). If a single-hit always results in infection, that is, *r* = 1, *Eq*.2 can be further simplified

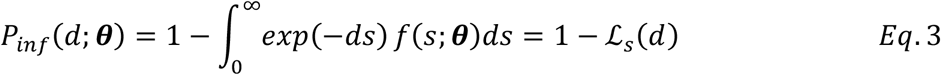

where ℒ_*s*_ refers to the Laplace transform of *f*(*s*).

As an illustrative example, we introduce the dose-response model where the level of susceptibility is distributed as a Gamma distribution, *s* ∼ Gamma(α, *β*). By solving *Eq*.3, the dose-response model is derived as

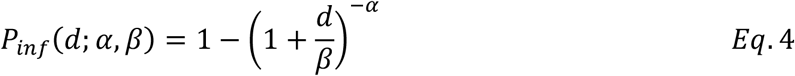

and this formula is the same as the so-called Beta-Poisson model [15,19]. Note that we can derive *E*q.4 without violating the single-hit principle, and the equation can be interpreted as the relationship between dose-dependent infection probability and the susceptibility distribution within a host.

#### Model fitting

Since the results of controlled infections are obtained as either infected or not in challenge experiments, such observation process leads to a binomial likelihood

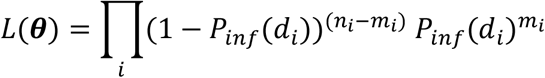

and thus the log-likelihood is

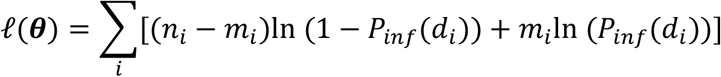

where for each trial *i* we have a dose *d*_*i*_ and a group of *n*_*i*_ volunteers of which *m*_*i*_ are infected. To estimate the set of parameters ***θ***, maximum likelihood estimation (MLE) was performed. For this computation we used the optim() function in the R statistical programming environment version 3.5.1., and 95 % confidence intervals were computed from 1000 bootstrapped samples.

#### Referred animal dose-response model

Current risk assessments of SARS-CoV-2 infection risk among humans often refer to the animal dose-response model obtained by Watanabe et al. [20]. Their study used a Delta model (i.e., the first model in **Table S3**) and fitted it to available SARS-CoV-1 data based on mouse experiments. As a result, the estimated parameter was 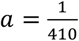 in **Table S3** notation. For details, see the original article [20]. For comparison of dose-response curves, we converted the unit of inoculated doses using the ratio of PFU to TCID_50_ that is previously established as 0.7 [21].

## Supplementary figures and tables

**Figure S1.**
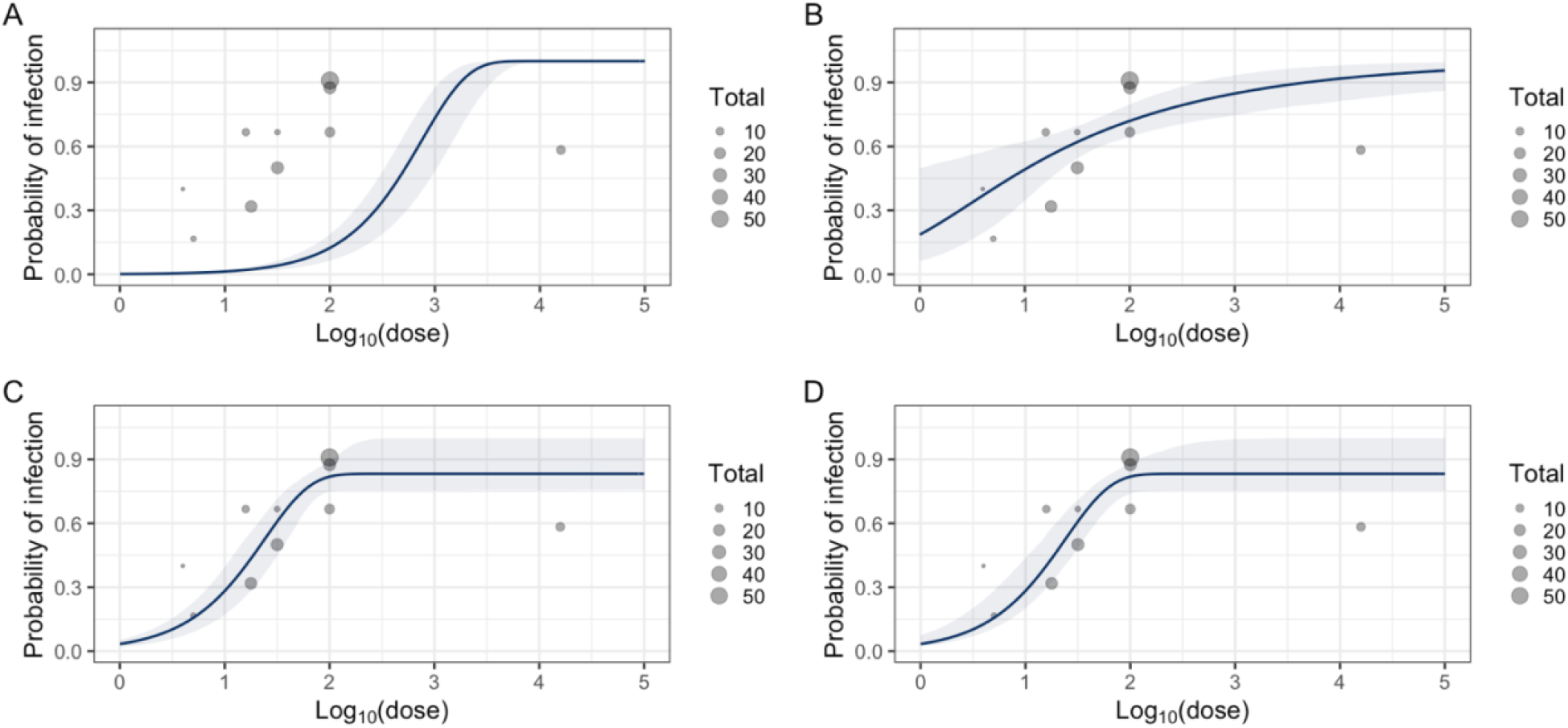
Estimated dose-response models of endemic coronavirus based on observed human challenge data. Grey bubbles indicate the observed data, and the size of bubbles indicates the number of participants in each trial. To describe different heterogeneity in susceptibility, dose-response models with Delta (panel **A**), Gamma (panel **B**), Two-level (panel **C**), and Gamma with point-mass distributions (panel **D**) were fitted to the data.

**Figure S2.**
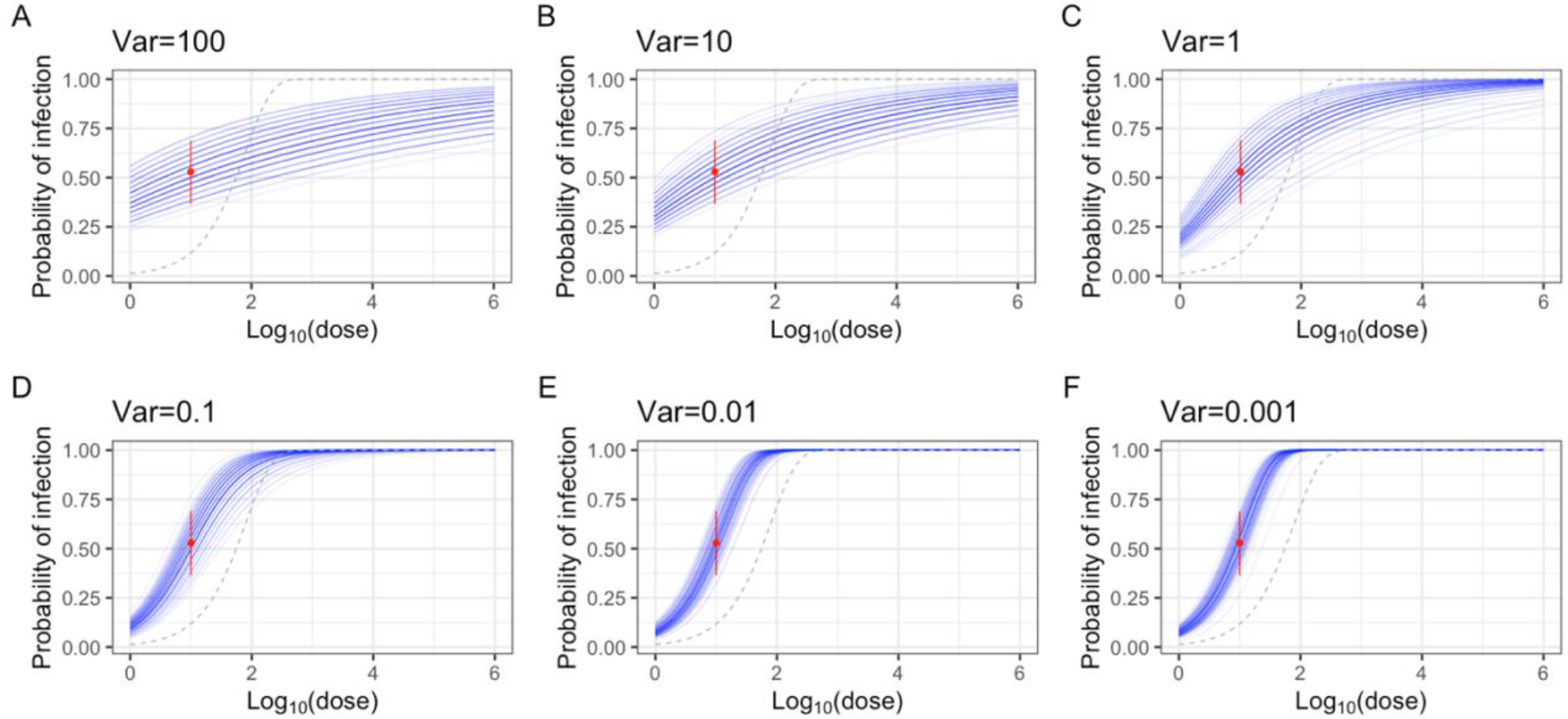
Simulated SARS-CoV-2 dose response curves based on observed human challenge data. Red plot and whiskers indicate the observed data and its 95% binomial confidence intervals. Each curve is obtained by bootstrapping with a gamma model. From panel **A** to **F**, the coefficient of variation is decreased from 10^2^ to 10^−3^.

**Table S1.** Human challenge data with endemic coronavirus.

| Dose | Total | Infected | Unit | Pathogen | Age | Inoculation | Reference |
| --- | --- | --- | --- | --- | --- | --- | --- |
| 100 | 24 | 21 | TCID50* | HCoV-229E | 21-53 | Nasal drop | Bende 1989 (Table 1) |
| 31.6 | 6 | 4 | TCID50 | HCoV-229E | 18-50 | Nasal drop | Bradburne 1967 (Fig 1) |
| 5.0 | 6 | 1 | TCID50 | HCoV-229E | 18-50 | Nasal drop | Bradburne 1967 (Fig 1) |
| 4.0 | 5 | 2 | TCID50 | HCoV-229E | 18-50 | Nasal drop | Bradburne 1967 (Fig 1) |
| 15.8 | 9 | 6 | TCID50 | HCoV-229E | 18-50 | Nasal drop | Bradburne 1967 (Fig 1) |
| 100 | 15 | 10 | TCID50 | HCoV-229E | Unknown | Nasal drop | Callow 1990 (Main text) |
| 17.8 | 22 | 7 | TCID50 | HCoV-229E | 18-50 | Nasal drop | Reed 1984 (Table V) |
| 31.6 | 24 | 12 | TCID50 | HCoV-229E | 18-50 | Nasal drop | Reed 1984 (Table V) |
| 15848.9 | 12 | 7 | TCID50 | HCoV-229E | 18-50 | Nasal drop | Reed 1984 (Table V) |
| 100 | 55 | 50 | TCID50 | HCoV-229E | 18-53 | Nasal drop | Tyrell 1993 (Table 2) |

\*TCID50: Median tissue culture infectious dose.

**Table S2.** Human challenge data with SARS-CoV-2.

| Dose | Total | Infected | Unit | Pathogen | Age | Inoculation | Reference |
| --- | --- | --- | --- | --- | --- | --- | --- |
| 10 | 34 | 18 | TCID50* | SARS-CoV-2<sup>†</sup> | 18-29 | Nasal drop | Killingley 2022 |

\*TCID50: Median tissue culture infectious dose.
<sup>†</sup>Pre-alpha wild-type virus (Genbank accession number OM294022).

**Table S3.** Description of dose-response models with different distributions for describing the heterogeneity in susceptibility against endemic coronavirus and its estimated parameters.

| Susceptibility distribution | Dose-response model | Estimated parameters | Description |
| --- | --- | --- | --- |
| Delta | $P_{inf}(d) = 1 - \exp(-ad)$ | $a = 1.3 \times 10^{-3}$<br>[$6.6 \times 10^{-4}, 2.3 \times 10^{-3}$] | All individuals have the same level of susceptibility, and the variance is zero. |
| Gamma | $P_{inf}(d) = 1 - (1 + \theta d)^{-k}$ | $\theta = 1.2$<br>[$1.4 \times 10^{-1}, 3.9 \times 10^2$]<br>$k = 2.7 \times 10^{-1}$<br>[$1.1 \times 10^{-1}, 5.3 \times 10^{-1}$] | The level of susceptibility for all individuals follows a gamma distribution. |
| Two-level | $P_{inf}(d) = 1 - [p_1 \exp(-a_1 d) + (1 - p_1) \exp(-a_2 d)]$ | $a_1 = 4.2 \times 10^{-9}$<br>[$5.8 \times 10^{-13}, 2.5 \times 10^{-5}$]<br>$a_2 = 4.2 \times 10^{-2}$<br>[$1.9 \times 10^{-2}, 6.9 \times 10^{-2}$]<br>$p_1 = 1.7 \times 10^{-1}$<br>[$1.4 \times 10^{-3}, 2.4 \times 10^{-1}$] | A population has two levels of susceptibility; a proportion ($p_1$) has a mass at level of susceptibility $a_1$ and the other has another mass at level of susceptibility $a_2$. |
| Gamma with point mass | $P_{inf}(d) = 1 - [p_1 \exp(-a_1 d) + (1 - p_1)(1 + \theta d)^{-k}]$ | $a_1 = 7.5 \times 10^{-17}$<br>[$9.7 \times 10^{-36}, 4.2 \times 10^{-5}$]<br>$p_1 = 1.7 \times 10^{-1}$<br>[$4.2 \times 10^{-7}, 2.5 \times 10^{-1}$]<br>$\theta = 6.2 \times 10^{-7}$<br>[$4.9 \times 10^{-8}, 1.0 \times 10^{-1}$]<br>$k = 6.8 \times 10^4$<br>[$7.4 \times 10^{-1}, 7.7 \times 10^5$] | The level of susceptibility for a proportion $1 - p_1$ of population follows gamma distribution, and the other proportion $p_1$ has a level of susceptibility $a_1$. |

**Table S4.** Comparison of estimated dose-response models based on model fit to the observed endemic coronavirus challenge data.

|  | Delta | Gamma | Two-level | Gamma+point-mass |
| --- | --- | --- | --- | --- |
| Log-Likelihood | -393.0 | -107.2 | -98.3 | -98.3 |
| No. of parameters | 1 | 2 | 3 | 4 |
| AIC* | 787.9 | 218.5 | 202.6 | 204.6 |
| Difference in AIC<sup>†</sup> | 585.3 | 15.9 | 0 | 2.0 |

\*AIC: Akaike Information Criterion. The lowest value indicates the best model in terms of prediction.
<sup>†</sup>Difference of >10 indicates strong evidence [22]. The values here suggest substantial support for the heterogeneity in susceptibility.

## Notes

### Competing Interest Statement

The authors have declared no competing interest.

### Author Declarations

The study used (or will use) ONLY openly available human data that were originally published in previous studies.

